# Multi-ancestry rare variant burden analysis of E3 ubiquitin ligase genes in Parkinson’s disease

**DOI:** 10.64898/2026.11.01.26366966

**Authors:** Karen L. Velásquez-Méndez, Daniel F. Santos-Gil, Maria Teresa Periñan, Konstantin Senkevich, Global Parkinson’s Genetic Program (GP2)

**Affiliations:** Facultad de Medicina, Universidad Nacional de Colombia, Bogotá, Colombia; Unidad de Trastornos del Movimiento, Servicio de Neurología, Instituto de Biomedicina de Sevilla, Hospital Universitario Virgen del Rocío/CSIC/Universidad de Sevilla, Seville, Spain; Centro de Investigación Biomédica en Red sobre Enfermedades Neurodegenerativas, Instituto de Salud Carlos III, Madrid, Spain; Center for Preventive Neurology, Wolfson Institute of Population Health, Faculty of Medicine and Dentistry, Queen Mary University of London, London, UK; The Neuro (Montreal Neurological Institute-Hospital), McGill University, Montreal, Quebec, Canada; Department of Neurology and Neurosurgery, McGill University, Montreal, QC, Canada; Department of Specialized Medicine, Division of Medical Genetics, McGill University Health Centre, Montreal, QC, Canada

## Abstract

E3 ubiquitin ligases regulate protein degradation through the ubiquitin-proteasome system and have been implicated in early-onset Parkinson’s disease (EOPD). We performed a multi-ancestry rare-variant burden analysis of seven E3 ubiquitin ligase genes (*HERC1, IRF2BPL, KMT2D, RAPSN, RNF168, RNF216, TRIM2*) using GP2 Release 11 WGS data from 17,928 PD patients (including 6,659 EOPD) and 8,977 controls across five major ancestry groups (African, Central Asian, East Asian, European, and Middle Eastern). We did not identify any association between gene based rare burden and PD, using the optimised sequence kernel association test (SKAT-O).

---

E3 ubiquitin ligases are central regulators of proteostasis, controlling the selective degradation of misfolded, aggregated, or damaged proteins through the ubiquitin-proteasome system (UPS) [1]. By conferring substrate specificity, these enzymes target proteins for proteasomal clearance, a process essential for neuronal homeostasis and survival [2]. Impaired E3 ligase function disrupts the clearance of toxic proteins, increases neuronal vulnerability, and contributes to the pathogenesis of neurodegenerative diseases [3]. In Parkinson’s disease (PD), UPS dysfunction is reflected by the accumulation of α-synuclein and ubiquitinated proteins within Lewy bodies [4].

Early-onset Parkinson’s disease (EOPD, age at onset ≤50 years) is more likely to be caused by rare variants in single genes than with late-onset disease [5], including *PRKN* and other protein homeostasis genes may be relevant to EOPD [6]. Several genes encoding E3 ubiquitin ligases have been implicated in both neurodevelopmental and neurodegenerative disorders [7,8]. In addition to the well-established role of *PRKN*, recent studies have suggested that rare variants in *HERC1, IRF2BPL, KMT2D, RAPSN, RNF168, RNF21*6, and *TRIM2* may also contribute to EOPD susceptibility [9–11]. However, these findings are based on relatively small studies and require replication in independent cohorts and diverse populations, as rare variant associations are particularly sensitive to cohort composition, sample size, and analytical methodology.

To evaluate these candidate genes, we performed a gene-based rare variant burden analysis using whole-genome sequencing (WGS) data from the Global Parkinson’s Genetics Program (GP2) Release 11 (10.5281/zenodo.17753486) [12]. The dataset included 17,928 PD patients (including 6,659 with EOPD) and 8,977 neurologically healthy controls (Table 1). Only ancestry groups with at least 500 cases and 500 controls were included in burden analyses. PD case-control analyses included African, Central Asian, East Asian, European, and Middle Eastern ancestry groups, whereas EOPD analyses were restricted to East Asian and European groups meeting this threshold. Rare (minor allele frequency [MAF] <0.01 in gnomAD) and ultrarare (MAF <0.001 in gnomAD) variants were classified into four functional categories: potentially functional, coding, predicted loss-of-function (LOF), and highly deleterious (CADD >20). Potentially functional variants included nonsynonymous, splicing, untranslated region [UTR], stop-gain, stop-loss, and synonymous variants predicted to affect splicing. Gene-based burden testing was performed using SKAT-O implemented in RVTests [13], adjusted for sex, age at recruitment, and the first five genetic principal components. This analysis includes burden and dispersion analysis, allowing for the presence of both deleterious and protective alleles. Multiple testing was controlled using the false discovery rate (FDR).

**Table 1.** Demographic characteristics of the study cohort.

| Ancestry | PD |  |  | EOPD |  |  | Controls |  |  |
|---|---|---|---|---|---|---|---|---|---|
|  | N | Age, mean ± SD (years) | Females, n (%) | N | Age, mean ± SD (years) | Females, n (%) | N | Age, mean ± SD (years) | Females, n (%) |
| AFR | 1,348<sup>*</sup> | 59.2 ± 10.5 | 366 (27.2%) | 310 | 43.3 ± 6.4 | 95 (30.6%) | 2,143<sup>*</sup> | 61.8 ± 10.2 | 918 (42.8%) |
| AJ | 846 | 58.7 ± 13.0 | 265 (31.3%) | 192 | 42.3 ± 6.4 | 57 (29.7%) | 268 | 68.3 ± 11.7 | 144 (53.7%) |
| AMR | 821 | 53.2 ± 13.2 | 315 (38.4%) | 366 | 40.7 ± 7.6 | 160 (43.7%) | 313 | 58 ± 11.1 | 191 (61.0%) |
| CAH | 229 | 47.1 ± 13.0 | 86 (37.6%) | 100 | 39.9 ± 7.9 | 40 (40.0%) | 123 | 62.7 ± 11.2 | 79 (64.2%) |
| CAS | 579<sup>*</sup> | 56.9 ± 11.1 | 329 (58.6%) | 124 | 40.1 ± 7.5 | 50 (40.3%) | 986<sup>*</sup> | 57.9 ± 10.2 | 573 (58.1%) |
| EAS | 1,462<sup>*</sup> | 60.5 ± 12.4 | 650 (44.5%) | 939<sup>†</sup> | 42.4 ± 6.8 | 443 (47.2%) | 1,057<sup>*†</sup> | 63.2 ± 10 | 537 (50.8%) |
| EUR | 11,551<sup>*</sup> | 54.7 ± 13.0 | 4,244 (36.7%) | 4,167<sup>†</sup> | 40.8 ± 7.3 | 1,587 (38.1%) | 2,761<sup>*†</sup> | 65 ± 12.8 | 1469 (53.2%) |
| MDE | 852<sup>*</sup> | 51.8 ± 11.2 | 375 (44.0%) | 257 | 41.5 ± 7.3 | 105 (40.9%) | 984<sup>*</sup> | 64.3 ± 14.5 | 564 (57.3%) |
| SAS | 240 | 56.4 ± 12.4 | 84 (35.0%) | 204 | 41.8 ± 6.6 | 57 (27.9%) | 342 | 57 ± 9.8 | 98 (28.7%) |
| *All cohorts* | 17,928 | 55.4 ± 12.9 | 6,714 (37.4%) | 6,659 | 41.2 ±7.2 | 2,594 (39.0%) | 8,977 | 62.8 ± 11.7 | 4573 (50.9%) |

*Abbreviations:* AFR, African; AJ, Ashkenazi Jewish; AMR, Admixed American; CAH, Complex Admixture History; CAS, Central Asian; EAS, East Asian; EUR, European; MDE, Middle Eastern; SAS, South Asian. Ancestries marked with an asterisk (\*) were included in the PD burden analyses, whereas those marked with a cross (†) were included in the EOPD burden analyses. Age is reported as age at recruitment for all groups. EOPD was defined as age at onset (AAO) ≤50 years.

In total, we identified 3,679 rare potentially functional variants, 1,549 coding variants, 9 predicted loss-of-function (LOF) variants, and 569 highly deleterious variants in the PD cohort (Supplementary Data 1). In the EOPD cohort, we identified 1,220 rare potentially functional variants, 555 coding variants, 3 predicted LOF variants, and 373 highly deleterious variants (Supplementary Data 2). Several variants were shared across ancestry groups in both cohorts, as summarized in Supplementary Figures S1–S5. Complete SKAT-O results are provided in Supplementary Data 4 and 5 for the PD and EOPD analyses, respectively.

No gene-based associations remained significant after FDR correction in either the PD or EOPD analyses. In the PD ultra-rare variant analysis (MAF<0.001), nominal associations were observed for *RAPSN* in the East Asian ancestry group (high-CADD; p=0.042, FDR=0.88) and *RNF168* in the European ancestry group (high-CADD; p=0.015, FDR=0.76).

In the EOPD rare variant analysis (MAF <0.01), nominal associations were found for *RAPSN* in the East Asian ancestry group (coding; p=0.048, FDR=0.30; high-CADD; p=0.046, FDR=0.30). In the EOPD ultrarare variant analysis (MAF <0.001), nominal associations were observed in the European group for *IRF2BPL* (coding; p=0.026, FDR=0.47; potentially functional; p=0.021, FDR=0.47), *RNF168* (coding: p=0.023, FDR=0.47), and *TRIM2* (high-CADD: p=0.028, FDR=0.47). These nominal findings are summarized in Tables 2 and 3 for the PD and EOPD analyses, respectively. No additional associations were identified for the remaining genes across ancestry groups, variant classes, or disease cohorts (Supplementary Data 4-5). No gene was nominally significant in more than one ancestry group.

**Table 2.**
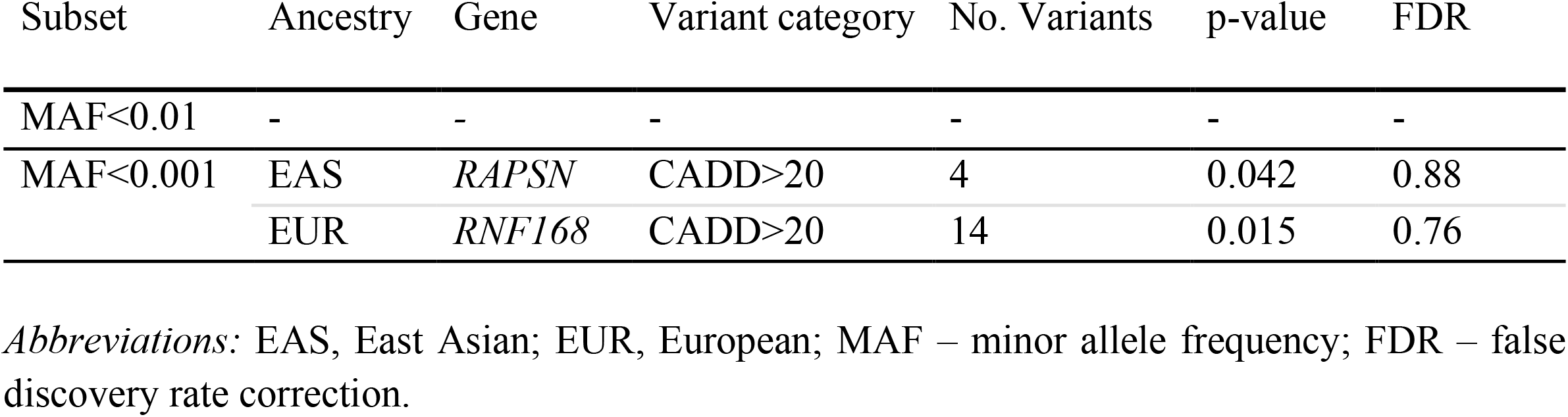
Summary of nominal burden associations identified in PD case-control analyses.

**Table 3.**
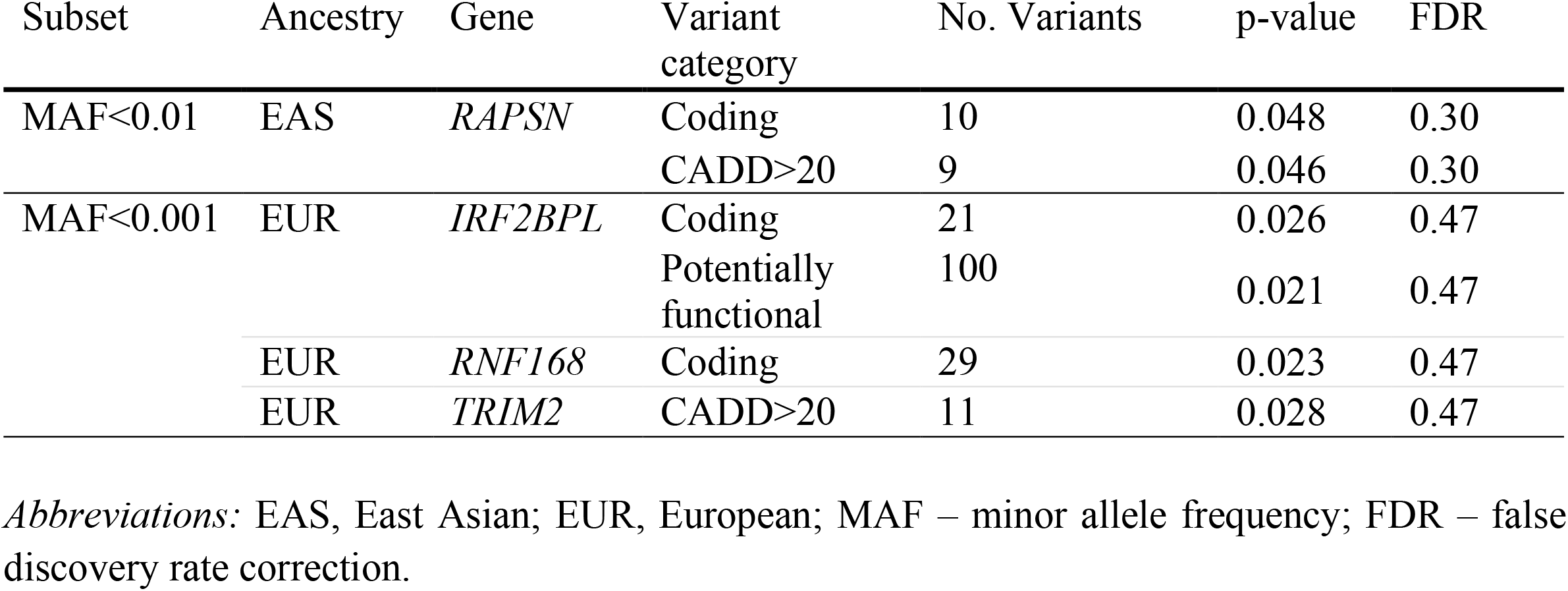
Summary of nominal burden associations identified in EOPD case-control analyses.

In this study, we performed a comprehensive, multi-ancestry, gene-based rare-variant burden analysis of seven candidate E3 ubiquitin ligase genes previously implicated in EOPD, using GP2 Release 11 WGS data. Although several nominal associations were identified across specific ancestry groups and variant categories, none remained significant after correction for multiple testing. Overall, our findings do not support a major contribution of rare variants in *HERC1, IRF2BPL, KMT2D, RAPSN, RNF168, RNF216*, and *TRIM2* to PD or EOPD susceptibility across ancestries.

The UPS plays a central role in protein homeostasis by degrading misfolded and aggregated proteins, including α-synuclein, and its dysfunction is well established in PD pathogenesis [1–5]. E3 ubiquitin ligases are particularly relevant within this pathway because they confer substrate specificity and regulate selective protein turnover [7,9,12]. Despite this strong biological rationale, we were unable to replicate previously reported associations [9–11,14]. Nominal associations were observed for *RAPSN*, primarily in individuals of East Asian ancestry, and for *RNF168, IRF2BPL*, and *TRIM2* in European and East Asian ancestry groups. However, these findings did not remain significant after multiple testing correction and should therefore be interpreted with caution. These observations may reflect chance findings, ancestry-specific variation, or limited carrier counts rather than true disease-associated effects.

Nevertheless, these genes remain biologically plausible candidates. *RAPSN* showed nominal associations in East Asian PD and EOPD analyses, particularly among highly deleterious variants. Although *RAPSN* is best known for its role in neuromuscular junction organization, emerging evidence suggests broader neuronal functions that may be relevant to neurodegeneration [9,15]. *RNF168*, which showed nominal associations in the European analyses, encodes an E3 ubiquitin ligase involved in the DNA damage response, a pathway increasingly implicated in neuronal aging and neurodegeneration [16]. *IRF2BPL* and *TRIM2* also showed nominal associations in ultra-rare variant analyses and have established roles in neuronal maintenance and neurodevelopment [11,17,18]. However, these findings should be interpreted cautiously until replicated in independent cohorts.

Some limitations should be acknowledged. Despite the large overall sample size, statistical power remains limited for rare and ultra-rare variants, particularly predicted LOF variants. In addition, several ancestry groups could not be included in the EOPD burden analyses due to insufficient sample sizes, thereby limiting the generalizability of these findings. Finally, these genes may contribute to PD susceptibility through mechanisms not captured in the present study, including regulatory variation, structural variants, epigenetic modifications, or gene-gene interactions.

Overall, our findings do not support a major contribution of rare variants in *HERC1, IRF2BPL, KMT2D, RAPSN, RNF168, RNF216*, and *TRIM2* to PD or EOPD susceptibility across ancestries. Although several nominal associations were observed, none remained significant after correction for multiple testing. These results do not rule out the role of the ubiquitin-proteasome system in PD pathogenesis; rather, they suggest that rare coding variation within these genes is unlikely to represent a major population-level contributor to PD or EOPD susceptibility.

## Methods

### Study cohort

We analyzed WGS data from the Global Parkinson’s Genetics Program (GP2) Release 11 [12]. The dataset included individuals with a confirmed diagnosis of PD and neurologically healthy controls. Demographic characteristics, including the number of cases and controls, mean age, standard deviation, and sex distribution, are summarized by ancestry group (Table 1). To ensure adequate statistical power for rare variant burden analyses, only ancestry groups with at least 500 PD patients and 500 controls were included. Power calculations indicated that ancestry groups below this threshold had <5% power to detect rare variant associations, supporting this inclusion criterion [19]. Accordingly, PD analyses included African, Central Asian, East Asian, European, and Middle Eastern ancestry groups. EOPD analyses (AAO ≤50 years) were restricted to East Asian and European ancestry groups meeting the same criterion. The final dataset comprised 17,928 PD patients, including 6,659 with EOPD, and 8,977 neurologically healthy controls (Table 1).

### Variant annotation and filtering

All preprocessing and downstream analyses were performed within the Verily® Workbench. Quality control procedures for the GP2 WGS dataset have been described previously [20]. In brief, the harmonized GP2 WGS pipeline retained variants meeting the following quality thresholds: genotype quality (GQ) ≥ 10, read depth (DP) ≥ 5, allele balance (AB) between 0.2 and

0.8 for heterozygous genotypes, and an overall variant call rate ≥ 0.95. Analyses focused on seven E3 ubiquitin ligase genes previously implicated in EOPD (*HERC1, IRF2BPL, KMT2D, RAPSN, RNF168, RNF216*, and *TRIM2*) and were performed separately within each ancestry group. Analyses were restricted to unrelated individuals by excluding second-degree or closer relatives. Population structure was accounted for using the first five genetic principal components (PC1-PC5).

Genomic regions corresponding to each candidate gene were extracted from the quality-controlled WGS data using PLINK and converted to Variant Call Format (VCF). Variants were annotated using ANNOVAR (hg38), utilizing resources from refGene, ClinVar, dbNSFP, and gnomAD (global and ancestry-specific frequencies). Rare variants were defined as having a minor allele frequency (MAF) < 0.01, and ultra-rare variants as having a MAF < 0.001, based on the gnomAD database. Variants were classified into four categories according to predicted functional consequence: (i) potentially functional variants, including nonsynonymous, splicing, untranslated region (UTR), stop-gain, stop-loss, and synonymous variants predicted to affect splicing; (ii) coding variants, comprising synonymous and nonsynonymous exonic variants; (iii) predicted loss-of-function (LOF) variants, including stop-gain, stop-loss, frameshift, and splicing variants; and (iv) highly deleterious variants, defined by a Combined Annotation Dependent Depletion (CADD) Phred score > 20.

### Gene-based burden testing

Gene-based rare-variant burden testing was performed using RVTests [13] with the Sequence Kernel Association Test - Optimal (SKAT-O). Models were adjusted for sex, age at recruitment, and the first five PCs. Analyses were performed separately for PD and EOPD and conducted independently within each distinct ancestry group. False discovery rate (FDR) correction was applied to account for multiple testing. All analyses were performed in Python and R within Verily® Workbench.

## Data availability

Data used in the preparation of this article were obtained from the Global Parkinson’s Genetics Program (GP2; https://gp2.org). Specifically, we used Tier 2 data from GP2 release 11 (DOI 10.5281/zenodo.17753486, release 11). All GP2 data is hosted in collaboration with the Accelerating Medicines Partnership in Parkinson’s disease and is available via application on the website (https://amp-pd.org). For up-to-date information on the study, visit https://www.amp-pd.org. Qualified researchers are encouraged to apply for direct access to the data through AMP-PD. The GP2 and AMP-PD datasets analyzed during the current study are available through AMP-PD (https://amp-pd.org).

## Code availability

All code generated for this article and the identifiers for all software programs and packages used are available on GitHub (https://github.com/GP2code/WGS-E3-PD-EOPD) and were given a persistent identifier via Zenodo (10.5281/zenodo.21681404).

## Acknowledgments

We would like to express our sincere gratitude to all participants and collaborators whose contributions made this study possible. Data (DOI 10.5281/zenodo.17753486, release 11) and/or code used in the preparation of this article were obtained from Global Parkinson’s Genetics Program (GP2; https://gp2.org), a research program of Aligning Science Across Parkinson’s (ASAP) (https://ror.org/03zj4c476). ASAP is managed by the Coalition for Aligning Science (CAS) (https://ror.org/00rf4ez40). The Michael J. Fox Foundation for Parkinson’s Research (https://ror.org/03arq3225) is the funding and strategic partner. For a complete list of GP2 members, see doi.org/10.5281/zenodo.7904831.

## Author contributions

KLV-M: Methodology, data curation, formal analysis, data interpretation, visualization, and writing—original draft preparation. DFS-G: Methodology, data curation, formal analysis, data interpretation, and writing—review and editing. MTP: Data interpretation and writing—review, and critical review of the final version of the manuscript. KS: Conceptualization, supervision, methodology, resources, project administration, data interpretation, and writing—review and editing. All authors read and approved the final version of the manuscript.

## Supplementary information

All supplementary materials for this manuscript are openly archived and available within our repository on Zenodo (DOI: 10.5281/zenodo.21681404). These materials include Supplementary Figures S1-S4 (summarizing the overlap of rare variants across ancestry groups in the PD cohort) and Supplementary Figure S5 (summarizing the overlap of rare variants across ancestry groups in the EOPD cohort), as well as Supplementary Data 1 and 2 (single-variant association analysis results) and Supplementary Data 3 and 4 (gene-based burden analysis results).

