## Supplementary Figures S1-S5 for "Multi-ancestry rare variant burden analysis of E3 ubiquitin ligase genes in Parkinson’s disease"

##### Supplementary Data

- **Supplementary Data 1.** Single-variant association analysis results in the Parkinson's disease (PD) cohort.
- **Supplementary Data 2.** Single-variant association analysis results in the early-onset Parkinson's disease (EOPD) cohort.
- **Supplementary Data 3.** Gene-based burden analysis results in the PD cohort (SKAT-O).
- **Supplementary Data 4.** Gene-based burden analysis results in the EOPD cohort (SKAT-O).

##### Supplementary Figures

- **Supplementary Figure S1.** Overlap of potentially functional rare variants across ancestry groups in the PD cohort.
- **Supplementary Figure S2.** Overlap of coding variants across ancestry groups in the PD cohort.
- **Supplementary Figure S3.** Overlap of variants with CADD > 20 across ancestry groups in the PD cohort.
- **Supplementary Figure S4.** Overlap of loss-of-function (LoF) variants across ancestry groups in the PD cohort.
- **Supplementary Figure S5.** Overlap of potentially functional, coding, high CADD (>20), and loss-of-function (LoF) rare variants across ancestry groups in the EOPD cohort.

### PD, POTENTIALLY FUNCTIONAL VARIANTS

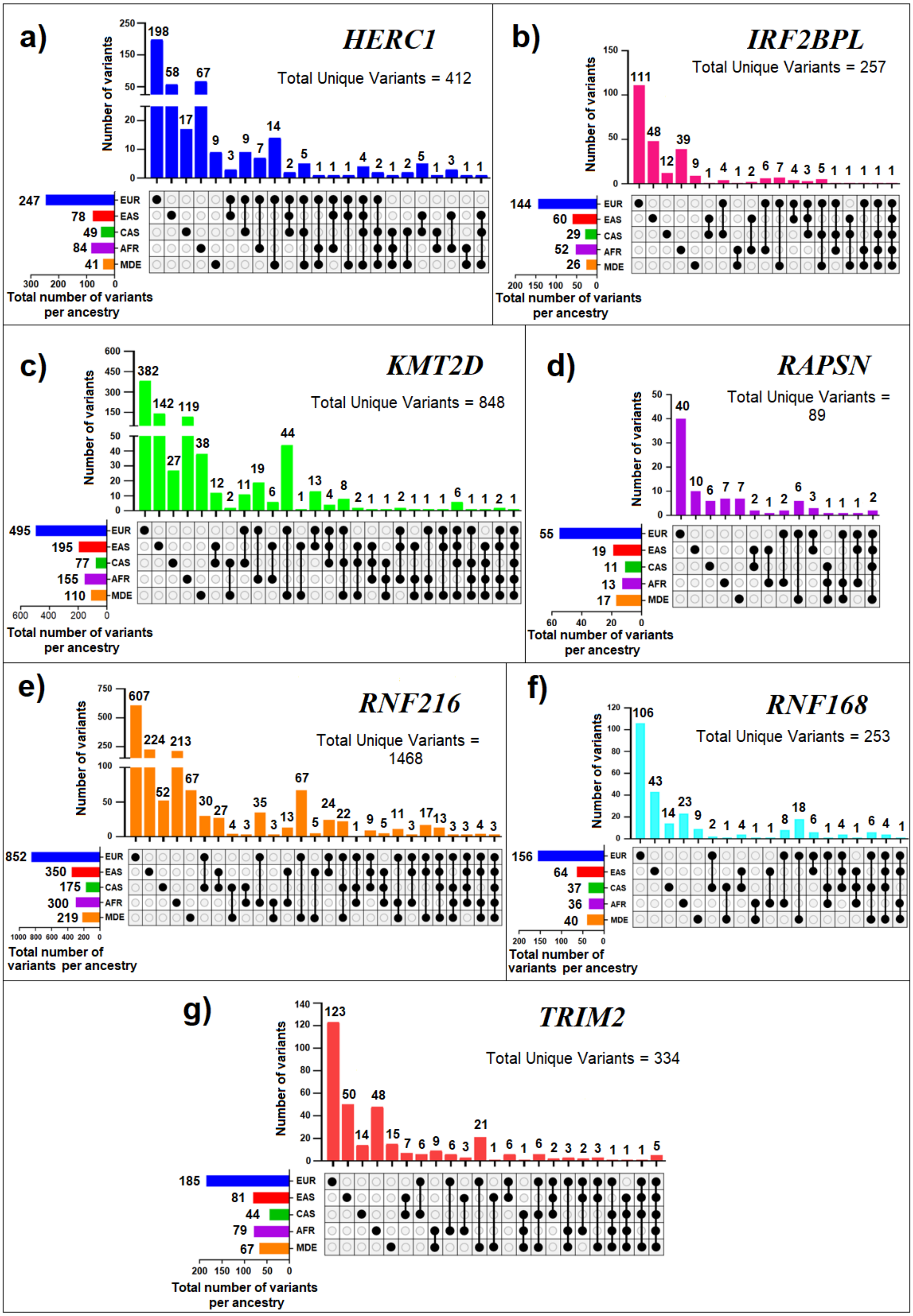

**Supplementary Figure S1. UpSet plots of potentially functional rare variants in the PD cohort.** Upper bars indicate the number of variants shared across ancestry combinations (EUR, EAS, CAS, AFR, and MDE), whereas left bars indicate the total number of variants identified in each ancestry. *Total Unique Variants* indicates the total number of unique variants identified for each gene across all ancestries. Panels correspond to (a) *HERC1*, (b) *IRF2BPL*, (c) *KMT2D*, (d) *RAPSN*, (e) *RNF216*, (f) *RNF168*, and (g) *TRIM2*.

### PD, CODING VARIANTS

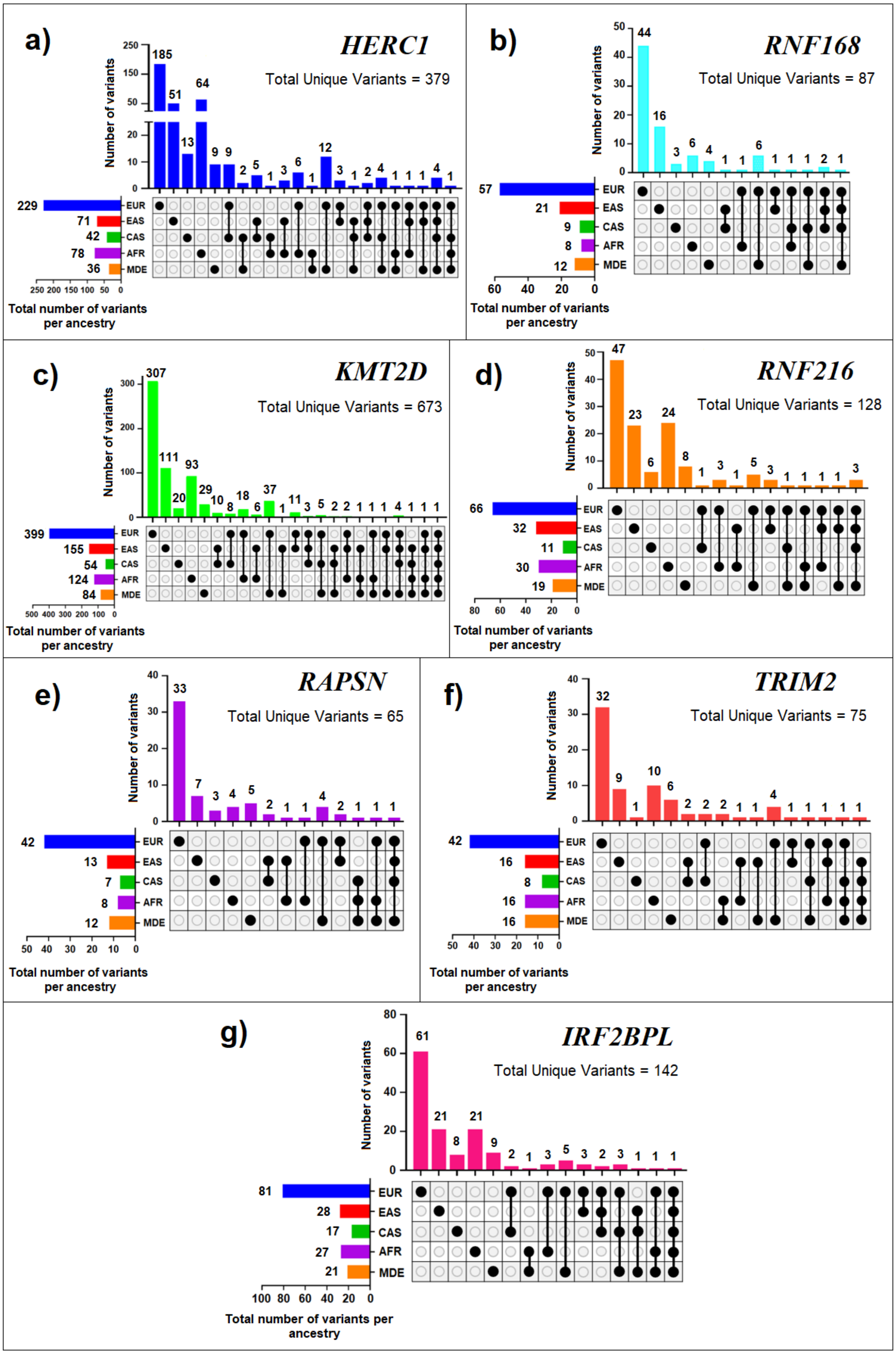

**Supplementary Figure S2. UpSet plots of coding rare variants in the PD cohort.** Upper bars indicate the number of variants shared across ancestry combinations (EUR, EAS, CAS, AFR, and MDE), whereas left bars indicate the total number of variants identified in each ancestry. *Total Unique Variants* indicates the total number of unique variants identified for each gene across all ancestries. Panels correspond to (a) *HERC1*, (b) *RNF168*, (c) *KMT2D*, (d) *RNF216*, (e) *RAPSN*, (f) *TRIM2*, and (g) *IRF2BPL*.

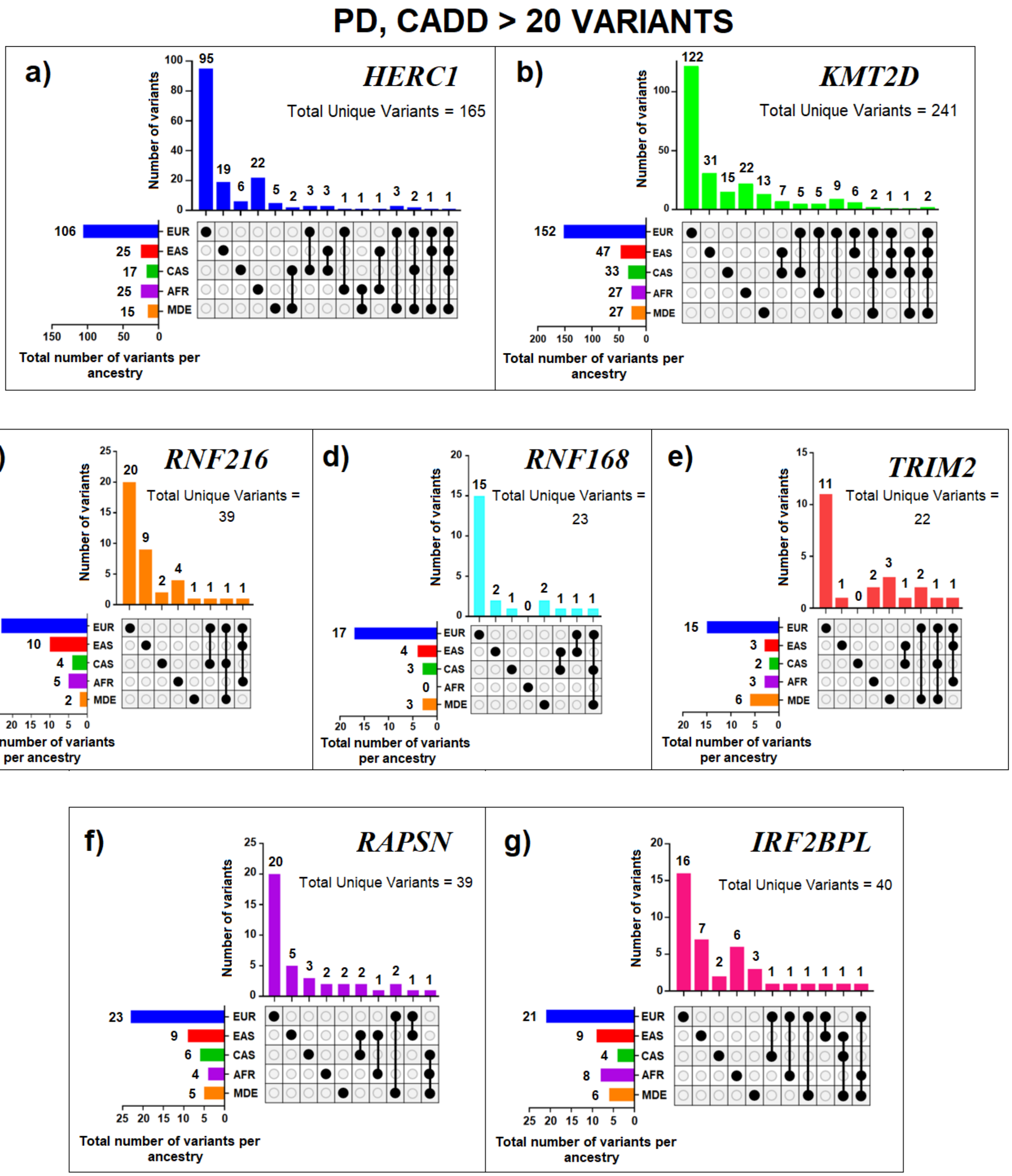


**Supplementary Figure S3. UpSet plots of CADD > 20 rare variants in the PD cohort.** Upper bars indicate the number of variants shared across ancestry combinations (EUR, EAS, CAS, AFR, and MDE), as indicated by the intersection matrix below. Left horizontal bars represent the total number of variants per ancestry. *Total Unique Variants* denotes the total number of unique variants identified for each gene across all ancestries. Only variants with CADD > 20 are shown. Panels correspond to (a) *HERC1*, (b) *KMT2D*, (c) *RNF216*, (d) *RNF168*, (e) *TRIM2*, (f) *RAPSN*, and (g) *IRF2BPL*.

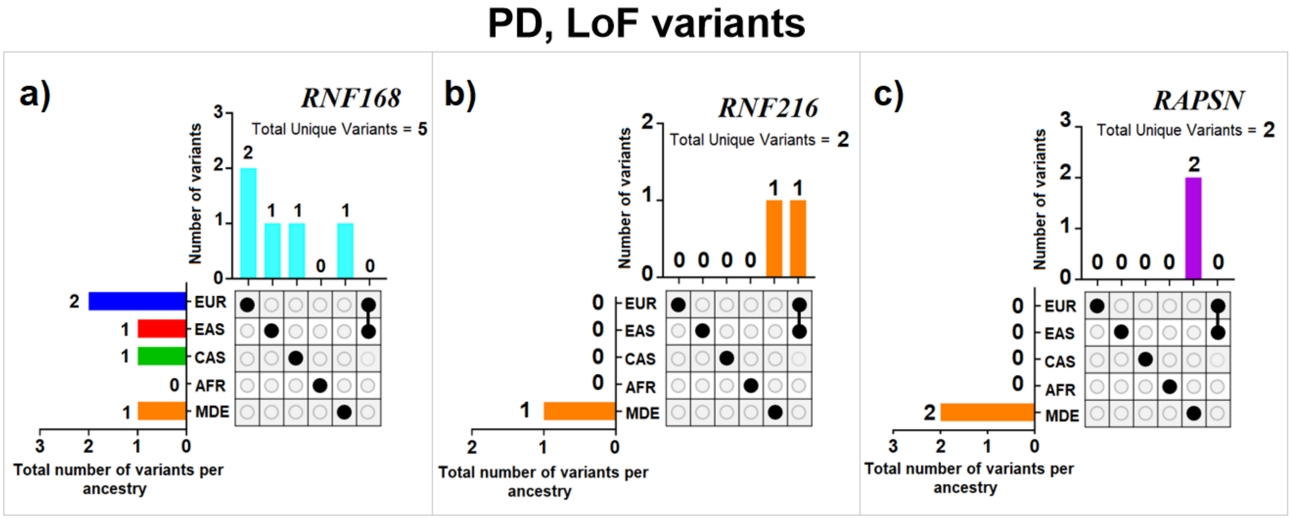


**Supplementary Figure S4. Loss-of-function (LoF) variants in the PD cohort.** Number of LoF variants identified across ancestry groups in the PD cohort. LoF variants were detected only in a) *RNF168*, b) *RNF216*, and c) *RAPSN*. No additional LoF variants were observed in the remaining genes analyzed.

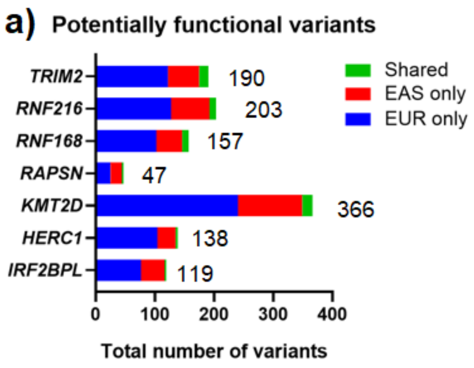


| Number of potentially functional variants in EOPD | | | |
|---|---|---|---|
| **Gene** | **EUR only** | **EAS only** | **Shared** |
| *TRIM2* | 122 | 53 | 15 |
| *RNF216* | 127 | 65 | 11 |
| *RNF168* | 102 | 44 | 11 |
| *RAPSN* | 25 | 19 | 3 |
| *KMT2D* | 240 | 109 | 17 |
| *HERC1* | 104 | 31 | 3 |
| *IRF2BPL* | 76 | 40 | 3 |

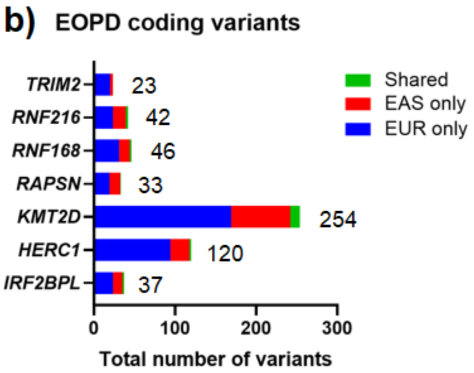


| Number of coding variants in EOPD | | | |
|---|---|---|---|
| **Gene** | **EUR only** | **EAS only** | **Shared** |
| *TRIM2* | 20 | 3 | 0 |
| *RNF216* | 23 | 16 | 3 |
| *RNF168* | 31 | 13 | 2 |
| *RAPSN* | 19 | 13 | 1 |
| *KMT2D* | 169 | 73 | 12 |
| *HERC1* | 94 | 24 | 2 |
| *IRF2BPL* | 23 | 12 | 2 |

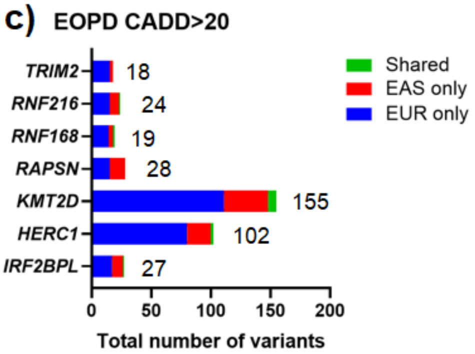


| Number of CADD>20 variants in EOPD | | | |
|---|---|---|---|
| **Gene** | **EUR only** | **EAS only** | **Shared** |
| *TRIM2* | 15 | 3 | 0 |
| *RNF216* | 15 | 8 | 1 |
| *RNF168* | 14 | 4 | 1 |
| *RAPSN* | 15 | 13 | 0 |
| *KMT2D* | 111 | 37 | 7 |
| *HERC1* | 80 | 20 | 2 |
| *IRF2BPL* | 17 | 9 | 1 |

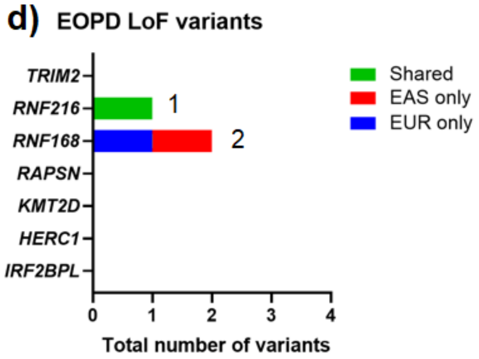


| Number of LoF variants in EOPD | | | |
|---|---|---|---|
| **Gene** | **EUR only** | **EAS only** | **Shared** |
| *TRIM2* | 0 | 0 | 0 |
| *RNF216* | 0 | 0 | 1 |
| *RNF168* | 1 | 1 | 0 |
| *RAPSN* | 0 | 0 | 0 |
| *KMT2D* | 0 | 0 | 0 |
| *HERC1* | 0 | 0 | 0 |
| *IRF2BPL* | 0 | 0 | 0 |

**Supplementary Figure S5. Distribution of variants across European (EUR) and East Asian (EAS) ancestries in the EOPD cohort for (a) potentially functional variants, (b) coding variants, (c) variants with CADD > 20, and d) LoF variants.** Stacked bars show the number of ancestry-specific variants (EUR only and EAS only) and variants shared between ancestries for each gene. Numbers beside the bars indicate the total number of variants per gene, while the accompanying tables provide the exact counts for each category.
